# Evaluating Mean Platelet Volume in relation to Disease Severity in Paediatric Sickle Cell Anaemia: A Cross-Sectional Study in Kwara State, North-Central Nigeria

**DOI:** 10.64898/2026.08.28.26361349

**Authors:** Oladimeji Faithful Damilola, Adewoyin Ademola David, Oyeleke Kikelomo

**Affiliations:** Department of Haematology and Blood Transfusion Science, College of Medicine, University of Lagos, Nigeria; Department of Medical Laboratory Services, Federal Neuropsychiatry Hospital, Budo-egba, Kwara State, Nigeria; Department of Medical Laboratory Science, Ladoke Akintola University of Technology, Ogbomoso, Nigeria

**Keywords:** Sickle cell anaemia, Mean platelet volume, Disease severity, Platelet indices, Paediatric haematology, Cross-sectional study, Nigeria

## Abstract

**Background:** Sickle cell anaemia (SCA) is characterised by chronic haemolysis, inflammation, platelet activation, and recurrent vaso-occlusive complications. Mean platelet volume (MPV) is a readily available platelet index, but evidence regarding its relationship with disease severity in paediatric SCA remains limited and inconsistent, particularly in African populations.

**Objective:** To evaluate the relationship between MPV and disease severity among children with SCA in Kwara State, North-Central Nigeria.

**Methods:** This hospital-based cross-sectional study included 51 clinically stable children with confirmed SCA consecutively recruited from the paediatric haematology clinic of Children Emergency Specialist Hospital, Ilorin. Complete blood count, including MPV, was performed using a Rayto RT-7600 automated haematology analyser. Disease severity was assessed using a composite clinical and laboratory scoring system based on a previously described method. Pearson’s correlation, Spearman’s rank correlation, simple linear regression, and the Kruskal–Wallis test were used as appropriate. Statistical significance was set at *p* < 0.05.

**Results:** Of 51 participants, 14 (27.5%) had mild, 33 (64.7%) moderate, and 4 (7.8%) severe disease. Mean MPV was 9.34 ± 0.76 fL (range, 8.0–11.2). Pearson’s correlation showed a weak positive, non-significant linear relationship with severity score (r = 0.231, *p* = 0.103), whereas Spearman’s analysis showed a weak positive monotonic association (ρ = 0.286, *p* = 0.042). Regression explained 5.3% of severity-score variation (R² = 0.053, *p* = 0.103). MPV did not differ significantly across severity categories (H = 2.163, *p* = 0.339). MPV correlated inversely with haemoglobin (r = −0.556, *p* < 0.001) and positively with platelet count (r = 0.307, *p* = 0.029).

**Conclusion:** MPV showed a weak relationship with disease severity but inconsistent statistical evidence across analyses. The limited explained variance and absence of significant differences between severity categories do not support MPV as a standalone severity marker. Larger longitudinal studies are warranted.

## 1.0 Introduction

Sickle cell disease (SCD) comprises a group of inherited haemoglobin disorders resulting from homozygous or compound heterozygous inheritance of pathogenic β-globin variants (1,2,3). Sickle cell anaemia (SCA), predominantly associated with the HbSS genotype, is the most common and clinically severe form of SCD and follows an autosomal recessive pattern of inheritance (2,4–6). A single nucleotide substitution in the β-globin gene (*HBB*) results in the replacement of glutamic acid by valine at the sixth amino acid position of the β-globin chain, producing haemoglobin S (HbS) (2,4,7–9). Under deoxygenated conditions, HbS polymerisation promotes erythrocyte sickling, chronic haemolysis, recurrent vaso-occlusion, endothelial dysfunction, inflammation, and progressive multisystem complications (2,10,11).

Sub-Saharan Africa bears a substantial proportion of the global burden of SCD, with large numbers of affected children born annually (12–16). Nigeria carries one of the largest burdens worldwide, with SCD affecting approximately 2-3% of the population (5,13,17,18). Despite improvements in supportive and preventive care, the clinical expression of SCA remains highly heterogeneous (3,10,19,20). Children may experience varying degrees of anaemia, vaso-occlusive complications, infection, acute chest syndrome, stroke, and other organ complications (7,19,21,22). This heterogeneity creates a need for simple and accessible laboratory parameters that may complement established clinical measures of disease severity, particularly in resource-constrained settings.

The pathophysiology of SCA involves chronic haemolysis, inflammation, oxidative stress, endothelial activation, and a persistent hypercoagulable state (10,23,24). Platelets contribute to these processes through interactions with erythrocytes, leucocytes, and vascular endothelial cells, potentially promoting vaso-occlusion and inflammatory responses (2,23,25,26). These processes may be accompanied by alterations in platelet indices, including mean platelet volume (MPV), platelet distribution width (PDW), plateletcrit (PCT), and platelet-large cell ratio (P-LCR) (27). MPV reflects the average volume of circulating platelets and is readily obtained as part of a routine complete blood count (28). Larger platelets are generally considered to be more metabolically and haemostatically active, providing a biological rationale for investigating MPV in disorders characterised by platelet activation, inflammation, and thrombosis (29).

Previous investigations have reported variable relationships between MPV and clinical characteristics or outcomes across disease state, and the SCD literature appears inconsistent (27,29–34). Some studies have reported higher MPV among patients with more severe disease or greater clinical activity, whereas others have found weak or non-significant associations (27,33,35,36). Differences in age, clinical status, disease severity definitions, treatment patterns, and population characteristics may contribute to these inconsistencies (14). Moreover, much of the available evidence has been derived from adult populations (5,33,34), while data specifically addressing paediatric SCA populations remain comparatively limited.

The evaluation of MPV in children with SCA is therefore of interest because it is inexpensive, routinely generated by automated haematology analysers, and does not require additional blood collection or specialised testing (28,37). Determining whether MPV is consistently associated with disease severity could clarify whether it has potential value as an adjunct to conventional clinical and laboratory assessment. However, an association observed in a cross-sectional study cannot establish prognostic value or predict future clinical outcomes. Therefore, this study evaluated the relationship between MPV and disease severity among clinically stable children with SCA attending a paediatric haematology clinic in Kwara State, North-Central Nigeria. We hypothesised that MPV would demonstrate an association with the clinical severity score.

## 2.0 Materials and Methods

### Study Design and Setting

This hospital-based analytical cross-sectional study was conducted at the Haematology Clinic of the Children Emergency Specialist Hospital, Centre Igboro, Ilorin, Kwara State, Nigeria. Ilorin is located between latitudes 8°30′ and 8°50′ North and longitudes 4°20′ and 4°35′ East, within the transitional ecological zone between Nigeria’s rainforest and Guinea savannah region of North-central Nigeria (38).The hospital provides specialised paediatric healthcare services, including routine follow-up care for children with sickle cell anaemia from Kwara State and surrounding communities.

### Study Population and Sampling

The study population comprised children and adolescents with confirmed sickle cell anaemia (HbSS genotype) attending the Haematology Clinic for routine follow-up care. Participants were recruited using consecutive sampling, whereby all eligible patients attending the clinic during the study period were considered for participation until the required sample size was reached. The minimum sample size was estimated using a formula for comparison of means between two independent groups, based on MPV estimates from a previous study (34). The previous study reported mean MPV values of 8.2 ± 0.5 fL among patients with fewer than three vaso-occlusive crises in the preceding year and 9.8 ± 1.1 fL among patients with more than three crises or an ongoing vaso-occlusive crisis.²⁶ Using a two-sided significance level of 5% and 90% statistical power, the estimated minimum sample size was approximately six participants per group. Given the small sample estimate and the anticipated heterogeneity of disease severity in the present paediatric cohort, a recruitment target of 48 participants was prespecified. Participants were recruited consecutively until this target was reached. A total of 51 eligible children were ultimately enrolled and included in the analysis.

### Eligibility Criteria Inclusion Criteria

Children and adolescents aged 2–17 years with confirmed sickle cell anaemia (HbSS genotype), established by cellulose acetate haemoglobin electrophoresis at alkaline pH (8.2–8.6), who were clinically stable at the time of recruitment and whose parents or legal guardians provided written informed consent were eligible for inclusion.

### Exclusion Criteria

Participants were excluded if they had haemoglobin variants other than HbSS, including HbSC, HbAS, or HbSβ-thalassaemia; were experiencing an acute vaso-occlusive crisis or other acute sickle cell-related complication at enrolment; had fever or clinically evident active infection at enrolment; were receiving corticosteroids, anticoagulants, or antiplatelet medications; or did not have written parental or legal guardian consent.

### Data Collection

Sociodemographic and relevant clinical information were obtained using a structured interviewer-administered questionnaire. Additional clinical information, including previous hospital admissions, transfusion history, and documented sickle cell-related complications, was obtained from participants’ medical records and case files for assessment of disease severity.

### Sample Collection and Processing

Following standard aseptic venepuncture procedures, 8 mL of venous blood was collected from the antecubital vein of each participant. Five millilitres were dispensed into ethylenediaminetetraacetic acid (EDTA) anticoagulant tubes for complete blood count analysis and haemoglobin genotype determination. Samples were processed within two hours of collection to minimise pre-analytical variation in haematological parameters, particularly MPV. The remaining 3 mL was collected into serum separator tubes for C-reactive protein (CRP) determination. Following clot formation and centrifugation, serum was separated and stored at −36°C until analysis.

### Laboratory Procedures Complete Blood Count

Complete blood count parameters, including platelet count and mean platelet volume, were measured using the Rayto RT-7600 automated haematology analyser (Rayto Life and Analytical Sciences Co., Ltd., Shenzhen, China), a 17-parameter, three-part differential analyser based on electrical impedance technology.

### Haemoglobin Genotype Determination

Haemoglobin genotype was determined using cellulose acetate electrophoresis at alkaline pH (8.2–8.6) following standard laboratory procedures. Haemoglobin variants were differentiated according to their electrophoretic mobility.

### Determination of C-Reactive Protein

Serum C-reactive protein concentrations were determined using a commercially available enzyme-linked immunosorbent assay (ELISA) kit (AccuBind ELISA System) according to the manufacturer’s instructions. All samples were analysed in duplicate at Shofalen Diagnostic Centre, Ilorin, Nigeria.

### Clinical Severity Assessment

Disease severity was assessed using a composite scoring system based on the method described by Manafa et al (39). The score incorporated haemoglobin concentration, white blood cell count, transfusion history, hospital admission frequency, and documented sickle cell-related complications. The anaemia component was scored according to haemoglobin concentration, while white blood cell count was categorised according to predefined ranges. Transfusion burden was calculated from the total number of transfusions relative to age, and hospital admission frequency was calculated from the number of admissions during the preceding three years. Documented sickle cell-related complications were also assigned scores according to the scoring framework. The resulting total severity score was used to categorise participants as having mild disease (score ≤3), moderate disease (score 4–7), or severe disease (score ≥8). The scoring framework was used as an operational measure of disease severity in the present study and was not considered a validated prognostic instrument.

### Statistical Analysis

Data were analysed using GraphPad Prism version 9 (GraphPad Software, San Diego, California, USA). Continuous variables were summarised using mean ± standard deviation (SD) for approximately normally distributed variables and median with interquartile range (IQR) for non-normally distributed variables. Categorical variables were presented as frequencies and percentages. The distribution of continuous variables was assessed using the Shapiro–Wilk test, supplemented by graphical inspection. The primary analysis evaluated the relationship between MPV and the continuous disease severity score. Pearson’s correlation coefficient was used to assess the linear association between MPV and severity score, while Spearman’s rank correlation coefficient was performed as a non-parametric sensitivity analysis because the severity score was not normally distributed. Simple linear regression was used to quantify the linear relationship between MPV and severity score, with severity score specified as the dependent variable and MPV as the independent variable. The regression coefficient, 95% confidence interval (CI), coefficient of determination (R²),

F statistic, and corresponding *p* value were reported. MPV distributions across the mild, moderate, and severe disease categories were compared using the Kruskal– Wallis test because of the non-normal distribution of the severity score and the small number of participants in the severe category. Because only four participants were classified as having severe disease, categorical comparisons were interpreted cautiously and considered exploratory. Pearson correlation analysis was also used to examine relationships between MPV and selected haematological and inflammatory parameters. All tests were two-tailed, and *p* < 0.05 was considered statistically significant.

### Ethical Considerations

Ethical approval was obtained from the Ethics and Research Committee of the Kwara State Ministry of Health, Ilorin, Nigeria (Reference No. MOH/KS/EU/777/593). Written informed consent was obtained from the parent or legal guardian of each participant before enrolment. Participant confidentiality was maintained by anonymising study records and restricting access to identifiable information. Participation was voluntary, and participants could withdraw from the study at any stage without prejudice to their clinical care.

## 3.0 Results

### Sociodemographic Characteristics

A total of 51 children with SCA were recruited. Participants were aged 2–17 years, with a mean age of 6.76 ± 4.39 years and a median age of 5 years. Thirty participants (58.8%) were male and 21 (41.2%) were female, giving a male-to-female ratio of 1.4:1.

### Disease Severity Classification

Fourteen participants (27.5%) had mild disease, 33 (64.7%) had moderate disease, and four (7.8%) had severe disease. The mean severity score was 4.63 ± 1.82, with scores ranging from 1 to 10. Moderate disease was therefore the predominant severity category.

### Mean Platelet Volume Analysis

MPV ranged from 8.0 to 11.2 fL, with an overall mean of 9.34 ± 0.76 fL. Across severity categories, mean MPV was 9.10 ± 0.75 fL in participants with mild disease, 9.43 ± 0.79 fL in those with moderate disease, and 9.43 ± 0.53 fL in those with severe disease. The corresponding median MPV values were 8.75 fL, 9.40 fL, and 9.25 fL, respectively.

### Correlation Between MPV and Disease Severity

**Table 1:** Correlation Analysis Between MPV and Clinical Severity Score.

| Variable | MPV (fL) | Severity Score |
| --- | --- | --- |
| MPV (fL) | 1.000 | 0.231 |
| *p-value* | - | 0.103 |
| Severity Score | 0.231 | 1.000 |
| *p-value* | 0.103 | - |
| N | 51 | 51 |

*Pearson correlation coefficient; two-tailed test*.

Pearson correlation demonstrated a weak positive but statistically non-significant linear association between MPV and disease severity score (r = 0.231, *p* = 0.103). Because the severity score was not normally distributed, Spearman’s rank correlation was also performed. This demonstrated a weak positive monotonic association between MPV and severity score (ρ = 0.286, 95% CI 0.002–0.526; *p* = 0.042).

### Regression Analysis

Simple linear regression, with severity score as the dependent variable and MPV as the independent variable, showed a positive but statistically non-significant regression coefficient (β = 0.582, 95% CI −0.123 to 1.286; *p* = 0.103). The model explained 5.3% of the variation in severity scores (R² = 0.053; F(1,49) = 2.755).

**Figure 1:**
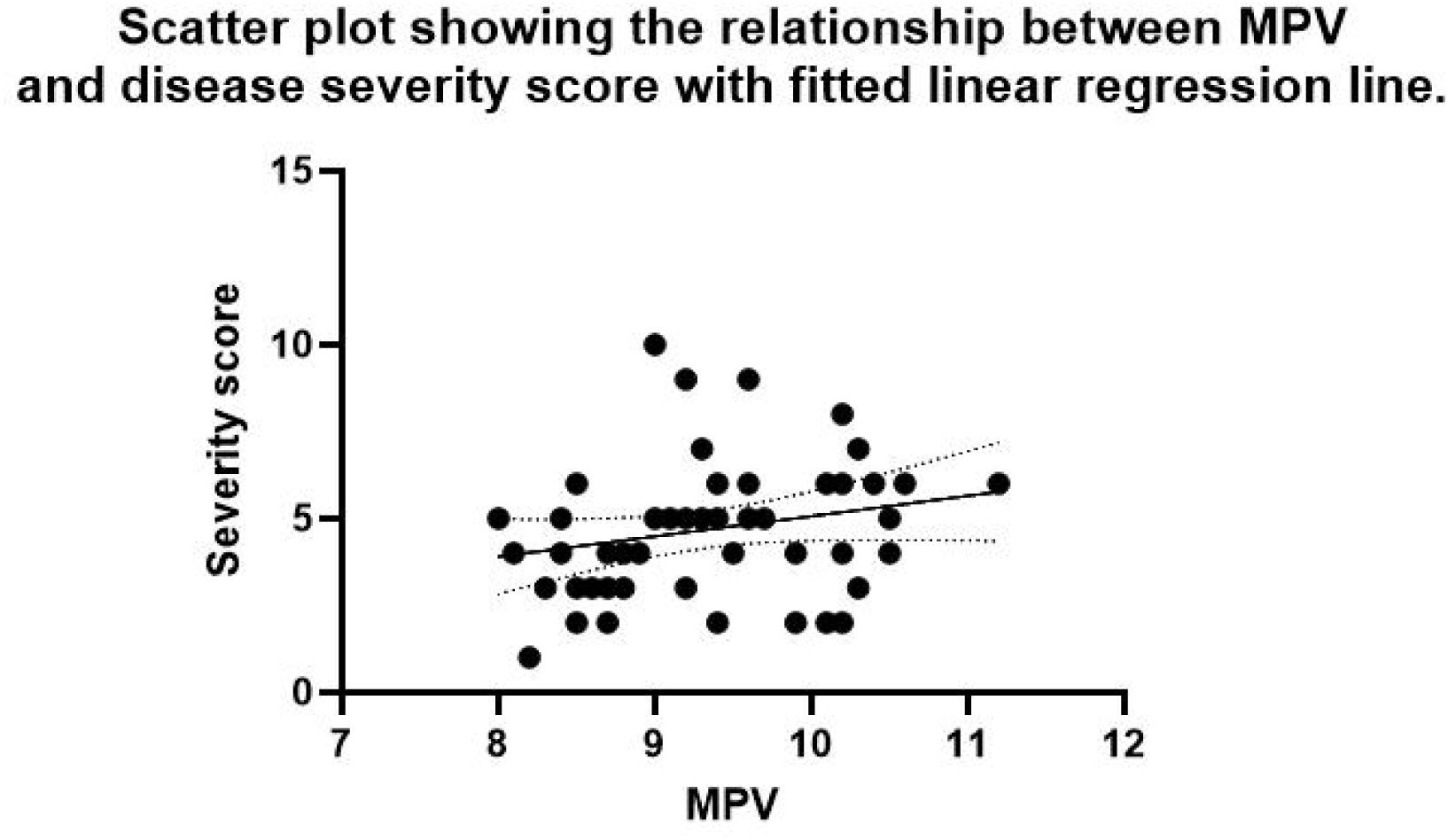
Linear Regression Analysis of MPV and Clinical Severity Score [Linear regression scatter plot showing MPV (y-axis, 8-12 fL) vs Severity Score (x-axis, 0-10) with regression line, 95% confidence intervals, and R² value displayed]

### MPV Across Disease Severity Categories

**Table 2.** Distribution of MPV according to disease severity category.

| **Disease severity** | **n** | **Mean ± SD (fL)** | **Median (IQR), fL** |
| --- | --- | --- | --- |
| Mild | 14 | 9.10 ± 0.75 | 8.75 (8.50–9.95) |
| Moderate | 33 | 9.43 ± 0.79 | 9.40 (8.85–10.15) |
| Severe | 4 | 9.43 ± 0.53 | 9.25 (9.05–9.98) |
| **Kruskal–Wallis H** |  | **2.163** |  |
| ***p* value** |  | **0.339** |  |

Kruskal–Wallis test; severe disease category comprised four participants.

The distribution of MPV did not differ significantly across the mild, moderate, and severe disease categories (Kruskal–Wallis H = 2.163, *p* = 0.339). Median MPV was 8.75 fL in the mild group, 9.40 fL in the moderate group, and 9.25 fL in the severe group.

### MPV and Other Haematological Parameters

**Table 3:** Correlation Analysis of MPV with Key Haematological Parameters.

| Parameter | Correlation Coefficient (r) | p-value | 95% CI | Interpretation |
| --- | --- | --- | --- | --- |
| Haemoglobin (g/dL) | -0.556 | 0.0001* | -0.71 to -0.35 | Strong negative |
| Platelet Count ($\times 10^9/L$) | 0.307 | 0.029* | 0.03 to 0.54 | Weak positive |
| White Blood Cell Count ($\times 10^9/L$) | 0.265 | 0.060 | -0.01 to 0.50 | Weak positive (NS) |
| Packed Cell Volume (%) | -0.102 | 0.476 | -0.37 to 0.18 | Weak negative (NS) |
| C-Reactive Protein (mg/L) | 0.230 | 0.109 | -0.05 to 0.48 | Weak positive (NS) |

*Significant at $p < 0.05$ ; NS = Not significant; CI = Confidence Interval*

MPV demonstrated a significant inverse correlation with haemoglobin concentration (r = −0.556, 95% CI −0.71 to −0.35; *p* < 0.001) and a weak positive correlation with platelet count (r = 0.307, 95% CI 0.03–0.54; *p* = 0.029). Correlations with white blood cell count (r = 0.265, *p* = 0.060), packed cell volume (r = −0.102, *p* = 0.476), and CRP (r = 0.230, *p* = 0.109) were not statistically significant.

## DISCUSSION

To our knowledge, this is among the few studies to examine the relationship between mean platelet volume (MPV) and disease severity specifically among clinically stable children with sickle cell anaemia (SCA)in Kwara State, North-Central Nigeria. MPV showed a weak positive relationship with severity score, although the findings varied according to the analytical method. Pearson correlation demonstrated a weak, non-significant linear association (*r* = 0.231, *p* = 0.103), whereas Spearman correlation showed a weak positive monotonic association that reached statistical significance (ρ = 0.286, *p* = 0.042). However, simple linear regression explained only 5.3% of the variation in severity score (R² = 0.0532, *p* = 0.103), and MPV did not differ significantly across mild, moderate, and severe categories (Kruskal–Wallis *H* = 2.163, *p* = 0.339). Collectively, these findings provide insufficient evidence to support MPV as a standalone marker of global disease severity in clinically stable children with SCA.

This finding is consistent with the multidimensional nature of SCA severity. Paediatric studies have more consistently associated disease severity with haemoglobin or packed cell volume, white blood cell count, fetal haemoglobin, reticulocyte burden, and clinical complications than with isolated platelet indices (40–44). Thus, a single platelet-size parameter such as MPV may not adequately capture the combined effects of anaemia, haemolysis, inflammation, vaso-occlusion, endothelial dysfunction, and cumulative organ involvement. Evidence from non-SCD paediatric populations similarly suggests that MPV may have greater value as an adjunctive rather than standalone marker of disease severity (45,46).

Nevertheless, opposing paediatric evidence indicates that MPV may be clinically informative under specific circumstances. In a Kenyan cohort of 104 children with SCD, MPV differed significantly across clinical phenotypes, with higher values reported in vaso-occlusive crisis and ischaemic stroke phenotypes (47). Higher MPV or related platelet indices have also been reported during crisis compared with steady state among Sudanese and Nigerian children (27,48,49). These findings do not necessarily contradict the present results because those studies primarily evaluated acute clinical states or specific complications, whereas our study assessed global severity among clinically stable children. Acute vaso-occlusive events may produce greater platelet activation and release of larger circulating platelets, making MPV more responsive to acute disease activity than to differences in baseline severity (27,50). Recent reviews similarly suggest that many SCD biomarkers discriminate vaso-occlusive episodes from steady state more readily than they predict chronic disease severity (51).

Our findings also differ from some adult SCA studies reporting higher MPV with greater disease severity. Khandekar et al. reported elevated MPV among adults with severe SCA compared with those with milder disease (34). Differences in age, disease duration, severity definitions, treatment exposure, and cumulative endothelial and organ injury may explain these differences. Adults may have greater chronic vasculopathy and thrombo-inflammatory burden, potentially strengthening associations between platelet activation and disease severity (24,52,53). In children, fetal haemoglobin and age-dependent biological factors may exert greater influence on disease expression and potentially attenuate relationships between individual platelet indices and global severity (40,54,55). The restriction of this study to clinically stable participants may also have reduced variation in MPV. This is supported by evidence showing greater abnormalities of platelet indices during vaso-occlusive or other acute events than during steady state (27,49,56). In addition, MPV is influenced by several processes beyond SCA severity, including inflammation, platelet production and consumption, bone marrow activity, and treatment exposure (57,58). Hydroxyurea may further modify the thrombo-inflammatory environment through effects on fetal haemoglobin, coagulation, and endothelial activation (59). These factors may partly obscure a cross-sectional relationship between MPV and global severity.

The strongest association observed was the inverse correlation between MPV and haemoglobin (*r* = −0.556, *p* < 0.001), indicating that lower haemoglobin concentrations were associated with larger platelets. Increased platelet turnover in chronic anaemia and haemolysis, with release of younger and larger platelets, provides a plausible explanation (24). African SCD data similarly demonstrate relationships between lower haemoglobin and increased platelet, leucocyte, and haemolytic burden (60), while studies of procoagulant vesicles have linked greater haemolysis with increased procoagulant activity (50,53). However, this finding requires particular caution because haemoglobin was itself a component of the composite severity score. Therefore, the weak MPV-severity relationship may partly reflect the stronger MPV-haemoglobin association rather than an independent relationship between MPV and overall disease severity. This is important because haemoglobin is independently associated with clinical outcomes in paediatric SCA (40). MPV also showed a weak positive correlation with platelet count (*r* = 0.307, *p* = 0.029). This may reflect the combined effects of chronic inflammation, haemolysis, functional hyposplenism, and increased platelet production or activation in SCA (61). However, an inverse MPV–platelet relationship has also been reported in SCD and other conditions, where increased platelet consumption may stimulate the release of larger immature platelets (62–64). Differences in age, clinical state, platelet turnover, and treatment may therefore account for the inconsistent direction of this association.

The mean MPV in this cohort (9.34 ± 0.76 fL) was comparable to values reported in some paediatric African and steady-state SCD populations, although higher values have been reported during crisis and in some adult cohorts (27,47,65). The predominance of moderate disease was broadly comparable with observations from other African and Nigerian paediatric cohorts, although differences in severity definitions limit direct comparison (41,43,44). The observed male predominance should not be interpreted as a sex-related difference in disease severity because sex associations have been inconsistent across paediatric African studies (41,42,44). Overall, the present findings do not support MPV as a standalone marker of disease severity in clinically stable children with SCA. Although the significant Spearman correlation suggests a weak monotonic relationship, the non-significant Pearson correlation, low explained variance in regression, and absence of significant differences across severity categories indicate limited clinical utility. Severity assessment is better supported by established clinical and laboratory parameters, including haemoglobin, fetal haemoglobin, white blood cell and reticulocyte measures, and complication history (40,41,44). MPV may nevertheless warrant further investigation as an adjunctive marker of acute disease activity or specific complications.

### Strengths and Limitations

Strengths of this study include the use of standardised laboratory procedures, consecutive recruitment of eligible participants, assessment of MPV using an automated haematology analyser, and evaluation of the MPV–severity relationship using complementary parametric and non-parametric statistical approaches. However, several limitations should be considered. First, the cross-sectional design precludes causal inference and assessment of the ability of MPV to predict future clinical events. Second, only clinically stable participants were enrolled, which may have restricted the range of MPV values and disease activity. Third, the sample size was relatively small, particularly in the severe disease category, which contained only four participants and limited the precision of subgroup comparisons. Fourth, the composite severity score incorporated haemoglobin concentration, which was itself significantly associated with MPV; this may have contributed to the observed relationship between MPV and the overall severity score. Fifth, fetal haemoglobin was not measured, despite its recognised relevance to disease phenotype. Finally, concurrent inflammatory conditions and other factors that may influence MPV could not be completely excluded.

### Future Directions

Future studies should employ longitudinal designs to assess MPV changes over time and in relation to clinical events. Studies including both steady-state and crisis episodes would provide more comprehensive assessment of MPV dynamics. Investigation of MPV alongside other emerging biomarkers, including fetal haemoglobin, soluble adhesion molecules, and inflammatory cytokines, may reveal multimarker panels with improved prognostic value. Additionally, research examining MPV in specific complication subtypes (e.g., stroke risk, acute chest syndrome) may identify niche applications for this parameter.

## Conclusion

Mean platelet volume demonstrated a weak positive relationship with disease severity among clinically stable children with sickle cell anaemia, although the statistical evidence varied according to the analytical approach. The linear association was not statistically significant and MPV explained only a small proportion of the variation in severity scores, while MPV distributions did not differ significantly across the predefined severity categories. Although MPV was significantly associated with haemoglobin concentration and platelet count, these findings do not establish MPV as a clinically reliable standalone marker of disease severity. Larger, adequately powered longitudinal studies are required to determine whether MPV provides clinically meaningful information beyond established measures of disease severity in paediatric SCA.

## Author Contributions

FDO: Conceptualization, data collection, laboratory analysis, statistical analysis, manuscript writing. AA: Supervision, methodology, manuscript review and editing. OK: Critical review and revision. All authors approved the final manuscript.

## Data Availability

All data produced in the present work are contained in the manuscript

## Acknowledgments

We thank the staff of the Haematology clinic, Children Emergency Specialist Hospital, Ilorin, and the patients and families who participated in this study.

## Data Availability

The datasets used in this study are available from the corresponding author upon reasonable request.

